# Machine learning identifies six genetic variants and alterations in the Heart Atrial Appendage as key contributors to PD risk predictivity

**DOI:** 10.1101/2021.06.29.21259734

**Authors:** Daniel Ho, William Schierding, Sophie Farrow, Antony A. Cooper, Andreas W. Kempa-Liehr, Justin M. O’Sullivan

## Abstract

Parkinson’s disease (PD) is a complex neurodegenerative disease with a range of causes and clinical presentations. Over 76 genetic loci (comprising 90 SNPs) have been associated with PD by the most recent GWAS meta-analysis. Most of these PD-associated variants are located in non-coding regions of the genome and it is difficult to understand what they are doing and how they contribute to the aetiology of PD. We hypothesised that PD-associated genetic variants modulate disease risk through tissue-specific expression quantitative trait loci (eQTL) effects. We developed and validated a machine learning approach that integrated tissue-specific eQTL data on known PD-associated genetic variants with PD case and control genotypes from the Wellcome Trust Case Control Consortium, the UK Biobank, and NeuroX. In so doing, our analysis ranked the tissue-specific transcription effects for PD-associated genetic variants and estimated their relative contributions to PD risk. We identified roles for SNPs that are connected with *INPP5P, CNTN1, GBA* and *SNCA* in PD. Ranking the variants and tissue-specific eQTL effects contributing most to the machine learning model suggested a key role in the risk of developing PD for two variants (rs7617877 and rs6808178) and eQTL associated transcriptional changes of *EAF1-AS1* within the heart atrial appendage. Similarly, effects associated with eQTLs located within the brain cerebellum were also recognized to confer major PD risk. These findings warrant further mechanistic investigations to determine if these transcriptional changes could act as early contributors to PD risk and disease development.

## Introduction

Parkinson’s disease (PD) is a complex neurodegenerative disease with a range of causes and clinical presentations. The diagnosis of PD is based on the presence of the cardinal motor symptoms (bradykinesia; muscular rigidity; 4-6 Hz resting tremor; postural instability)^1^. Genome wide association studies (GWAS) have identified human genetic variants that are associated with the risk of developing PD^2,3^. There are 290 PD-associated GWAS SNPs listed in the GWAS catalog. In the most recent PD GWAS meta-analysis, *Nalls et al*. identified 90 independent single nucleotide polymorphisms (SNPs) that are significantly associated with PD risk^2^. However, it is difficult to understand how these variants confer PD risk because the majority of the PD SNPs are located in non-coding regions of the genome^4–6^.

Non-coding SNPs have been shown to be enriched at regulatory loci and can act as expression quantitative trait loci (eQTLs)^7–11^. eQTLs typically explain a fraction of the variation in mRNA expression levels for target genes, either in *cis* (<1Mb apart in the linear sequence) or *trans* (>1Mb apart or located on a different chromosome). Regulatory variants (i.e. eQTLs) can impact different genes in different tissues, making it challenging to determine how SNPs convey risk for a phenotype. Determining the relative contributions of the eQTLs to the risk of developing a disease would help identify the eQTL-gene-tissue combinations that convey the risk associated with the variant. We have demonstrated that the three-dimensional structure of the genome can be used to help identify eQTL-gene pairs and thus the biological pathways that putatively contribute to disease etiology^12,13^. Yet, approaches that calculate relative estimates of the tissue specific contributions that SNPs make to disease development remain elusive.

We reasoned that if PD-associated SNPs contribute to disease development through gene regulatory effects, then the tissue-specificity of these eQTLs may be an important consideration for the aetiology of the disease^14,15^. Therefore, we developed a machine-learning predictor model for PD disease status that utilises and selects SNPs (without eQTLs in GTEx) and tissue-specific eQTL data, for case and control cohorts, to reveal the tissue-specific regulatory effects that are associated with PD risk. Briefly, we used a matrix of: 1) PD-associated SNPs that act as eQTLs, 2) the genes regulated by these eQTLs; 3) the tissues in which the eQTL effects were observed; and 4) SNPs that do not have eQTLs in GTEx to build a logistic predictor that was validated using genotype data from three independent studies^3,16,17^. The logistic predictor model that had the highest PD predictive ability, was trained and selected using the Wellcome Trust Case Control Consortium (WTCCC) cohort. The predictor model was then validated using two datasets derived from the UK Biobank^17^ and NeuroX-dbGap^16^. Our predictor ranked the relative contributions of six non-eQTL PD SNPs, and additional eQTLs that modulated gene regulation specifically within the heart atrial appendage as making the largest contributions to PD risk development.

## Methods

### Workflow for developing the PD predictor

We developed a machine learning approach that incorporates feature selection and cross validation, to calculate the additive tissue-specific contributions of spatial eQTLs within genotypes from individuals who developed PD (Figure 1).

**Figure 1.**
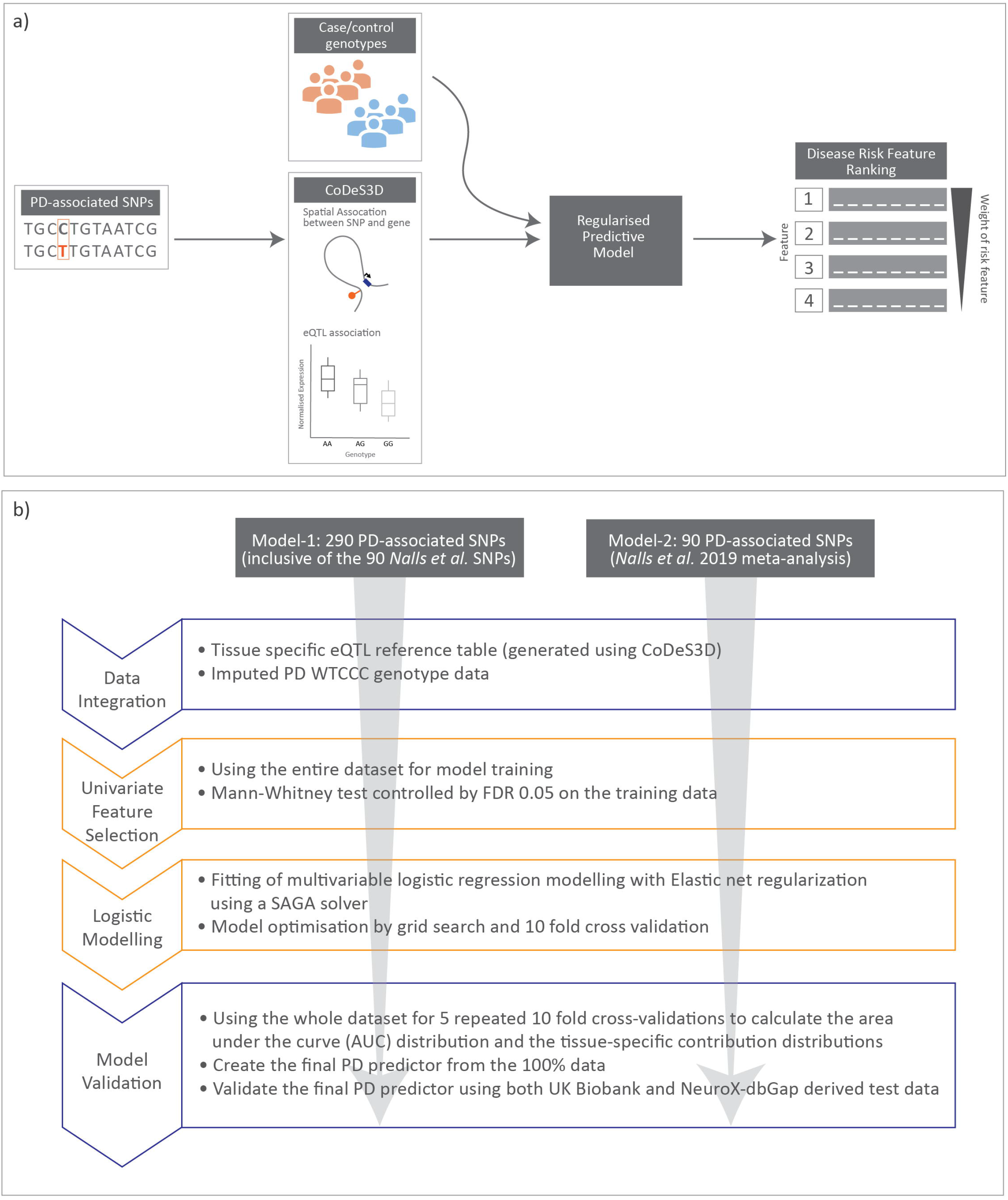
Cartoon illustrating data integration and workflow for regularised logistic regression modelling undertaken in this manuscript. a) Schematic diagram for data integration used to rank disease risk features. b) Workflow used to create the two regularised logistic regression predictor models for PD.

### Generation of tissue specific PD eQTL reference table

GWAS SNPs associated with PD (n=290, *p-*value <1.0 × 10^−5^; Supplementary Table 1) were obtained from the GWAS catalogue (www.ebi.ac.uk/gwas, downloaded 27^th^ August 2020). This SNP set included young adult-onset Parkinsonism SNPs^18^ and the 90 SNPs identified by the most recent meta-analysis by *Nalls et al*.^2^.

The PD-associated SNPs (Supplementary Table 1) were analysed using the Contextualize Developmental SNPs in 3-Dimensions (CoDeS3D) algorithm^6^, with the beta effect calculation option, to identify: a) the genes that physically interact with the PD-associated SNPs; and b) which of these SNP-gene interactions are eQTLs. Physical interactions between PD-associated SNPs and genes were identified using Hi-C chromatin contact libraries (Supplementary Table 2) captured from:

1. Cell lines from primary human tissues (*e*.*g*. brain, skin and spinal cord)
2. Immortalised cell lines that represent the embryonic germ layers (*i*.*e*. HUVEC, NHEK, HeLa, HMEC, IMR90, KBM7, K562, and GM12878)

These libraries were chosen to represent tissues with the largest number of possible interactions that occur in the human system.

The potential regulatory effects (normalized effect size [NES]) of the spatial connections were mapped by leveraging the eQTL information from 49 human tissues (Genotype-Tissue Expression database [GTEx] v8; www.gtexportal.org). eQTL significance levels were adjusted for multiple testing [Benjamini–Hochberg FDR]^19^ and considered significant if *q*<0.05.

### WTCCC cohort cleaning and genotype imputation

The PD genotype dataset was acquired from the WTCCC (Request ID 10584). The WTCCC PD genotype dataset, generated using Illumina microarrays, contained one case cohort (2197 individual samples) and two control cohorts (58C: 2930 individual samples and NBS: 2737 individual samples). Thus, the total number of control samples (n=5667) was more than double the number of cases. It is likely that the use of an imbalanced training dataset would create a biased disease status predictor. Therefore, only the control samples for the 58C 1958, British Birth cohort were used in this study.

SNPs and individual samples that were of poor quality and were recommended for study exclusion by the WTCCC were removed (Supplementary Table 3). SNPs within individual genotypes were converted to dbSNP rsIDs and genomic positions mapped (GRCh37, hg19) by Python scripts. PLINK (v1.90b6.2, 64-bit)^20^ was used for quality control. Genotypes were cleaned using the Method-of-moments F coefficient estimate to remove case homozygosity outliers (F values < -0.02 or 0.02 < F values) and the control outliers (F values < -0.016 or 0.19 < F values). Related individuals were identified and removed using proportion IBD (PI_HAT > 0.08). Ancestry outliers (identified by principal component analysis [PCA] plotting), individuals with sex genotype errors (identified by PLINK), or individuals with missing genotype data (missing rate > 5%) were also removed. Finally, SNPs that were significantly outside of Hardy-Weinberg Equilibrium (p < 10^−6^) or had a minor allele frequency < 1% were also removed.

The WTCCC PD case and control genotype data were obtained using two different Illumina microarrays (Human670-QuadCustom and Human1-2M-DuoCustom_v1_A)^3^. Therefore, we only used the 526,576 SNPs that were present in both microarrays for imputation. SNP data imputation was performed to recover a total of 27,590,399 SNPs using the Sanger imputation service (https://imputation.sanger.ac.uk), EAGLE+PBWT pipeline^21,22^, and Haplotype Reference Consortium(r1.1)^23^. Imputation was performed according to the default instructions (https://imputation.sanger.ac.uk/?instructions=1). Following imputation, PLINK was used to update the genotype data with rsIDs and remove SNPs with an: impute2 score < 0.3; missing data rate > 5%; or those that were not in Hardy-Weinberg Equilibrium (p < 10^−6^). The genotypes for 281 of the 290 PD SNPs used in this study (Supplementary Table 1) were extracted from the imputed PD genotype data.

### Creation of a weighted WTCCC PD genotype eQTL matrix

We created a matrix that combined individual genotypes with the eQTL effects for the PD-associated SNPs. There were three groups of data fields in the PD genotype eQTL table:

1. Individual sample information (sex, and disease status)
2. Individual sample PD-associated SNP genotype (SNP minor allele count) weighted by GTEx tissue-specific eQTL normalised effect sizes
3. Individual PD-associated SNP genotype for the SNPs with no eQTL effect information

The tissue-specific eQTL normalised effect size (NES) for the PD-associated SNPs were extracted from the GTEx eQTL summary table of significant eQTLs (Supplementary Table 4). The NES for each tissue-specific eQTL was weighted by the number of alternative alleles (0, 1 or 2) at the eQTL SNP position in each individual’s genome. 54 of the 290 PD-associated SNPs had no identifiable eQTL effects (Supplementary Table 5) and were input into the model unweighted, using solely SNP allele count from the imputed genotype.

We created two regularised logistic regression models (see below): for model-1, we created a weighted WTCCC PD genotype eQTL matrix for all 290 SNPs that were imputed and represented in the PD eQTL reference table. By contrast, for model-2 we created a weighted WTCCC PD genotype eQTL matrix for the subset of 90 SNPs from the PRS identified in *Nalls et al*.^2^.

### Generation, training, and validation of the regularised logistic regression models (model-1 and model-2)

We developed a regularised logistic regression predictor that incorporated a: 1) Mann-Whitney U tests in combination with Benjamini-Yekutieli procedure for controlling False Discovery Rate (FDR) to identify relevant information; and 2) multivariate prediction step that considers all features in context and removes redundant information, to identify the best combination of features for prediction of PD. Regularised logistic regression was incorporated into the models to enable the identification of features that contribute most to the final score.

The weighted WTCCC PD genotype eQTL matrix that contained all the case and control genotypes that passed quality control (4366 individual samples: 1698 cases and 2668 controls) was used to train model-1 (using all 290 SNPs), or model-2 (using the 90 *Nalls et al*. SNPs).

The Mann-Whitney U test (tsfresh version 0.16.0)^24^ was used to select the individual feature columns within the weighted WTCCC PD genotype eQTL matrix from the full training dataset that were the most relevant attributes for predicting PD status (*i*.*e*. the relevant attribute subset; FDR = 0.05)^25^. The relevant attribute subset was then used to train a multivariate logistic regression model (Scikit-learn version 0.23.2)^26^ implemented with elastic net regularisation using the SAGA solver to predict PD disease status. The machine learning elastic net regularisation prevented overfitting the predictor model by further sub-selecting the essential features for delivering the best prediction.

Training was optimised (measured by area under the receiver operating characteristic curve [AUC])^27^ using a Scikit-learn Grid Search algorithm^26,28^ with 10-fold cross-validation setting to select the optimised predictor model hyperparameters from the training stage (90% of cohort used for training, 10% used for cross validation). The optimised predictor hyperparameters for model-1 were: C=0.5, l1_ratio=0.6, max_iter=800, penalty=‘elasticnet’, random_state=1, solver=‘saga’ from the search space of following:

- ‘C’: 0.01, 0.05, 0.1, 0.5,1, 10, 20, 30,
- ‘max_iter’: 200, 500, 800, 1000, 1200, 1400, 1500, 1600,
- ‘l1_ratio’: 1, 0.9, 0.8, 0.7, 0.6, 0.5, 0.4, 0.3, 0.2, 0.1.

The optimised predictor hyperparameters for model-2 were: C=0.6, l1_ratio=0.1, max_iter=130, penalty=‘elasticnet’, random_state=1, solver=‘saga’ from the search of following:

- ‘C’: 0.0001, 0.001, 0.01, 0.1,0.2,0.3,0.4,0.5,0.6,0.7,0.8, 1, 3,
- ‘max_iter’: 1, 5, 70, 100, 130, 150, 170, 180, 200, 300, 500, 1000, 1200, 1400, 1600, 1800, 2000, 2200, 2400, 2600, 3000,
- ‘l1_ratio’: 1, 0.9, 0.8, 0.7, 0.6, 0.5, 0.4, 0.3, 0.2, 0.1.

The search space of models-1 and 2 did not include l1_ratio = 0 for excluding L2 regularisation and implementing feature selection. To calculate the variation in AUCs of the models with the optimised parameters we undertook 5 repeats of 10-fold cross-validation of model generation and validation by the Scikit-learn RepeatedKFold algorithm^26^. The 10-fold cross-validation started with the random generation of 10 equal parts from the full dataset. Nine parts of the data were used for training, and the remaining data were for validation. Mann Whitney U test filtering controlled by FDR = 0.05 was applied to the training set. Subsequently, the filtered training data were modelled by the multiple regularised logistic regression algorithm with the optimised predictor hyperparameters of (model-1 or model-2). This process was repeated until all parts of the data were used for validation. The result of this process is the final PD predictor for each model, with in-sample (training data) predictive performance as assessed by AUC.

### Calculation of tissue-specific contributions to PD risk

The 50 PD regularised logistic regression predictors created from the 5 repeats of 10-fold cross-validation above were used to test the predictive power of the model created with the optimised predictor hyperparameters from the tissue-specific eQTL effects. Tissue-specific contributions to the PD risk were extracted from each of the 50 PD regularised logistic regression predictors as the sum of the absolute values of the model weights associated with each tissue.

### Validation of model-1 and model-2

The generalising PD predictive power of models-1 and -2 was validated by testing on two independent test datasets derived from the UK Biobank and NeuroX-dbGap genotype data.

As the UK Biobank only has a small number of PD case samples, we created 30 different test cohorts of individual samples (without missing data for those SNPs included in model-1 or 2) using the same PD cases with 30 independently and randomly chosen non-PD diagnosed controls. Each cohort was used to create a weighted eQTL-genotype matrix for testing the predictive power and evaluating by AUC. The mean AUC of the 30 predictive tests was used as the validation result of the models.

NeuroX-dbGap was the largest PD single array study^2,16,29^, and we selected all the PD case and control samples of NeuroX-dbGap (without missing data for those SNPs included in model-1 or 2) to build a weighted eQTL-genotype matrix for validating the PD predictive power of each model.

Model-1 was validated using the following two independent datasets: 1) 30 cohorts of 2384 individual samples (928 cases and 1456 controls) derived from the UK Biobank; and 2) the 5,224 cases and 5,563 controls from NeuroX-dbGap (the largest PD single array study)^2,16,29^.

Model-2 was also validated by the same two independent datasets: 1) the 30 cohorts of 3812 individual samples (1484 cases and 2319 controls) derived from the UK Biobank; and 2) the cases and control from the NeuroX-dbGap dataset (5,224 cases and 5,563 controls) ^2,16,29^. Note the number of cases and controls in the UK Biobank differed as there were fewer cases excluded due to missing SNP data (see below).

### UK Biobank cohort definition and genotype imputation

Genotypes (case and control) that were used from the UK Biobank were selected as follows. European Caucasian samples identified by genetic clustering methods were selected and imputed (487,411 individual samples). The genomic relatedness analysis excluded SNPs that the UK Biobank recommended were removed from the selected case and control data.

The cases (model-1: 928 cases or model-2:1484 cases) were selected using the following criteria:

1. PD patient identified by the UK Biobank developed algorithm (field 42033)
2. PD patient identified by hospital records G20
3. PD patient had no missing data for any SNPs within the predictor model (model-1 or model-2). The greater number of SNPs used in model-1 meant that more cases were excluded due to missing data.

Control genotypes, not having records of Parkinsonism and without missing data for any of the SNPs included in the final predictor, were randomly selected from the healthy controls within the UK Biobank data for each of the 30 test cohorts. As model 2 had more cases, more controls were also included so as to match the ratio of case:control.

Genotype data of the UK Biobank case and control samples in each test cohort were used to build a weighted eQTL-genotype matrix for testing model-1 or 2 to recognise the disease status of individual samples correctly.

### NeuroX-dbGap cohort definition and genotype imputation

Genotypes were also obtained from the NeuroX-dbGap dataset. Genotypes were cleaned by removing all insertion and deletion variants. SNP IDs were converted to dbSNP rsIDs. Variants in chromosome 24(Y), 25(XY) and 26(MT) that are not included in the study due to the inconsistency with the Sanger imputed SNP data were also removed. Ancestry outliers (identified by principal component analysis [PCA] plotting), individuals with sex genotype errors (identified by PLINK), or individuals with missing genotype data (missing rate > 5%) were also removed. Finally, SNPs that were not in Hardy-Weinberg Equilibrium (p < 10^−5^) or had a minor allele frequency < 1% were removed. All the variants in the final model (model-1 or model-2) which were not present in the NeuroX-dbGap data were replaced with proxy SNPs using linkage disequilibrium information (r^2^> 0.5)^29^ calculated by PLINK from European 1000 genome genotype data (https://www.internationalgenome.org/about)^30^. The European 1000 genome genotype data were downloaded from (https://ctg.cncr.nl/software/magma)^31^ on 10^th^ August 2020.

### Mann-Whitney U test filtering on 290 PD and 313 T1D SNPs derived eQTL matrix

We generated a GTEx eQTL summary table of significant eQTLs (Supplementary Table 7) using 313 Type 1 Diabetes (T1D) associated SNPs (Supplementary Table 6). We mixed the 290 PD and 313 T1D SNPs to create a weighted WTCCC PD and T1D genotype eQTL matrix, as outlined above. Mann-Whitney U test filtering (controlled by FDR = 0.05) was applied to the weighted WTCCC PD and T1D genotype eQTL matrix to determine the filtering power for removing non-related PD features.

### Data analysis

All statistical tests were performed with Scikit-learn (version 0.23.2)^26^, and tsfresh (version 0.16.0)^24^

### Code Availability

The CoDeS3D pipeline is available at: https://github.com/Genome3d/codes3d-v2.

The Python scripts and machine learning code used in this analysis are available at: https://github.com/Genome3d/PD_lg_predictor_analysis

Python version 3.7.3 was used for all the Python scripts.

### Data availability

The data that support the findings of this study are available were derived from the following resources available in the public domain:

The SNPs are listed in Supplementary table 1.

The HiC datasets are listed in Supplementary table 2.

eQTL information from 49 human tissues were obtained from the Genotype-Tissue Expression database [GTEx] v8; www.gtexportal.org

PD genotype datasets were acquired from the: Wellcome Trust Case Control Consortium (Request ID 10584); UKBioBank (Application Number 61507); NeuroX-dbGap (dbGap project#98581-1)

## Results

### PD-associated SNPs are tissue specific eQTLs for 1,334 eGenes

We hypothesised that PD SNPs modulate disease risk through tissue-specific eQTL effects (*i*.*e*. eQTL-eGene)^14,15^. We analysed 290 PD-associated GWAS SNPs (Supplementary Table 8) for spatial eQTL interactions^7,32,33^ across 49 GTEx tissues^14^. 231 of the 290 (79.7%) PD SNPs tested were involved in 18,041 tissue-specific eQTL associations (Benjamini– Hochberg FDR < 0.05^19^; Supplementary Table 4), regulating 1,334 eGenes across the 49 GTEx tissues. Gene ontology analysis (David Functional Annotation)^34^ identified that the regulated genes were significantly enriched for intracellular signal transduction, antigen processing and presentation of peptides, among other pathways (Supplementary Table 9).

### Modelling genotype data to identify the genetic risk associated with tissue-specific eQTL effects for PD disease status

Understanding the impacts and complex networks associated with eQTLs is challenging. We hypothesised that regularised logistic regression models could be used to identify and rank the tissue-specific eQTLs that were significant contributors to PD risk.

We integrated the CoDeS3D eQTL analysis of the 290 PD SNPs with the genotype data for individuals within the WTCCC^35^ PD cohort (4366 individual samples: 1698 cases and 2668 controls; methods)^3^. Of the 290 PD SNPs, 281 SNPs were present in the WTCCC data. This resulted in the generation of a PD-SNP derived weighted WTCCC PD genotype eQTL effect matrix containing 17,829 tissue-specific eQTL-eGene pairs (227 SNPs, 1310 eGenes, 49 tissues) and 54 (of the 281) SNPs that had no known eQTL effects following our CoDeS3D analysis. Uninformative features for PD prediction were removed using a Mann-Whitney U test^36^ (FDR < 0.05) (Methods). After filtering, 11,288 PD SNP derived features (53 SNPs, 245 eGenes, 49 tissues) remained within the relevant attribute subset of the weighted WTCCC PD genotype eQTL effect matrix.

To test the effectiveness of the Mann-Whitney U test filter, we generated a PD and type 1 diabetes (T1D) SNP derived eQTL effect matrix using a mixed set of 290 PD and 313 T1D-associated SNPs and integrating with the WTCCC PD cohort genotypes (Supplementary Table 6). The PD + T1D SNP derived tissue-specific eQTL effect matrix included 25,052 SNP related data fields (556 SNPs, 1927 eGenes, 49 tissues). After the Mann-Whitney U test filtering (FDR < 0.05), 11,147 of the data fields (45 SNPs, 209 eGenes, 49 tissues) were selected using PD as the phenotypic outcome. Only one of the 313 (0.32%) T1D-associated SNP, rs1052553, remained following the Mann-Whitney U test filtering. Although rs1052553 has not previously been associated with PD in GWA studies, it has been implicated in PD as part of a PD risk haplotype^37,38^. Therefore, these results confirm that the Mann-Whitney U test filters uninformative data while preserving valuable PD information for our modelling.

We created regularised logistic regression models for PD risk using the Mann-Whitney U test filtered PD variant derived eQTL effect matrix (11,288 PD-SNP derived features [53 SNPs, 245 eGenes, 49 tissues]). The AUCs of the 50 PD regularised logistic regression predictors had a mean of 0.565 (distributed from 0.516 to 0.637) and a standard deviation of 0.024 (generated with the optimised predictor model hyperparameters by 5 repeats of 10-fold cross validation). The final PD predictor model (model-1) was trained using the entire WTCCC PD cohort. After the Mann-Whitney U test filtered WTCCC PD variant derived eQTL effect matrix contained 17,829 variant derived features. Model-1 selected 827 tissue-specific eQTLs and 6 SNPs with no eQTL effect (Supplementary Table 10). Model-1 had an enhanced diagnostic ability as represented by an AUC of 0.627 obtained using the training data.

We validated the predictive power of model-1 using two independent PD cohorts (UK Biobank^17^ (30 datasets of 923 cases and 1456 controls) and NeuroX-dbGap^16,29^). Model-1 was validated in both cohorts, producing mean AUCs of 0.572 and 0.571 in the UK BioBank and NeuroX-dbGap cohorts, respectively. These two validation results are highly consistent and within the range of the model AUCs (0.516 to 0.637) estimated by the 50 optimised logistic regression predictor models.

### eQTLs specific to the heart atrial appendage contribute to genetic risk in PD

We used the magnitude of the model weights (coefficients) for the genetic features, grouped by tissue-specificity of the effects, in the logistic regression model-1 as proxies for the contribution of the features to PD risk.

Six SNPs that had no identified eQTL effects (from CoDeS3D analysis of GTEx) made the most significant group contribution (18% of the total model weight) to the risk of PD development (Table 1; Figure 2). The six non-eQTL SNPs are: rs117896735, rs144210190, rs35749011, rs12726330, rs356220 and rs5019538 (Table 1). Note that the GTEx study^12^ removed rs356220 and rs5019538 from the tissue-specific eQTL data as part of their QC processing. Therefore, we were unable to test if rs356220 and rs5019538 were eQTLs. rs117896735 also has no eQTL effect information found in GTEx database. The other three SNPs (rs144210190, rs35749011 and rs12726330) were not detected by CoDeS3D to have spatial eQTL and eGene interactions within the Hi-C libraries used in this study.

**Table 1:** SNPs identified as being the main contributors to model-1. a) SNPs with no detected eQTL effects, and b) eQTL effects within the Heart Atrial Appendage. The model weight is the coefficient assigned to each variant or eQTL in the logistic regression predictor model-1. ‘^*^’ indicates the non eQTL SNP is in the 90 SNPs of *Nalls et al*.

| **SNP**<br>**(no detected eQTLs)** | **model weight** |
| --- | --- |
| *rs117896735_A | 0.42436 |
| rs1442190_A | 0.40106 |
| *rs35749011_A | 0.24949 |
| rs12726330_A | 0.18701 |
| rs356220_T | 0.17507 |
| *rs5019538_G | -0.08418 |

b)
| **Tisssue** | **eQTL**<br>**(rsID_major allele)** | **Gene** | **model weight** |
| --- | --- | --- | --- |
| Heart_Atrial_Appendage | rs7617877_A | EAF1-AS1 | 0.28996 |
| Heart_Atrial_Appendage | rs6808178_T | EAF1-AS1 | 0.25261 |
| Heart_Atrial_Appendage | rs6808178_T | TMEM161B-AS1 | 0.16339 |
| Heart_Atrial_Appendage | rs11707416_A | P2RY12 | 0.01163 |
| Heart_Atrial_Appendage | rs26431_G | EIF3KP1 | 0.0089 |
| Heart_Atrial_Appendage | rs17577094_G | RP11-259G18.3 | 0.00703 |
| Heart_Atrial_Appendage | rs365825_G | RP11-259G18.3 | -0.00467 |
| Heart_Atrial_Appendage | rs17577094_G | LRRC37A4P | -0.00434 |
| Heart_Atrial_Appendage | rs17577094_G | KANSL1-AS1 | 0.0036 |
| Heart_Atrial_Appendage | rs8070723_G | RP11-259G18.3 | -0.00294 |
| Heart_Atrial_Appendage | rs365825_G | LRRC37A4P | 0.00237 |
| Heart_Atrial_Appendage | rs365825_G | KANSL1-AS1 | -0.00128 |
| Heart_Atrial_Appendage | rs17577094_G | DND1P1 | 0.00116 |
| Heart_Atrial_Appendage | rs17577094_G | MAPK8IP1P2 | 0.00058 |
| Heart_Atrial_Appendage | rs199515_G | RP11-259G18.3 | -0.00011 |

**Figure 2:**
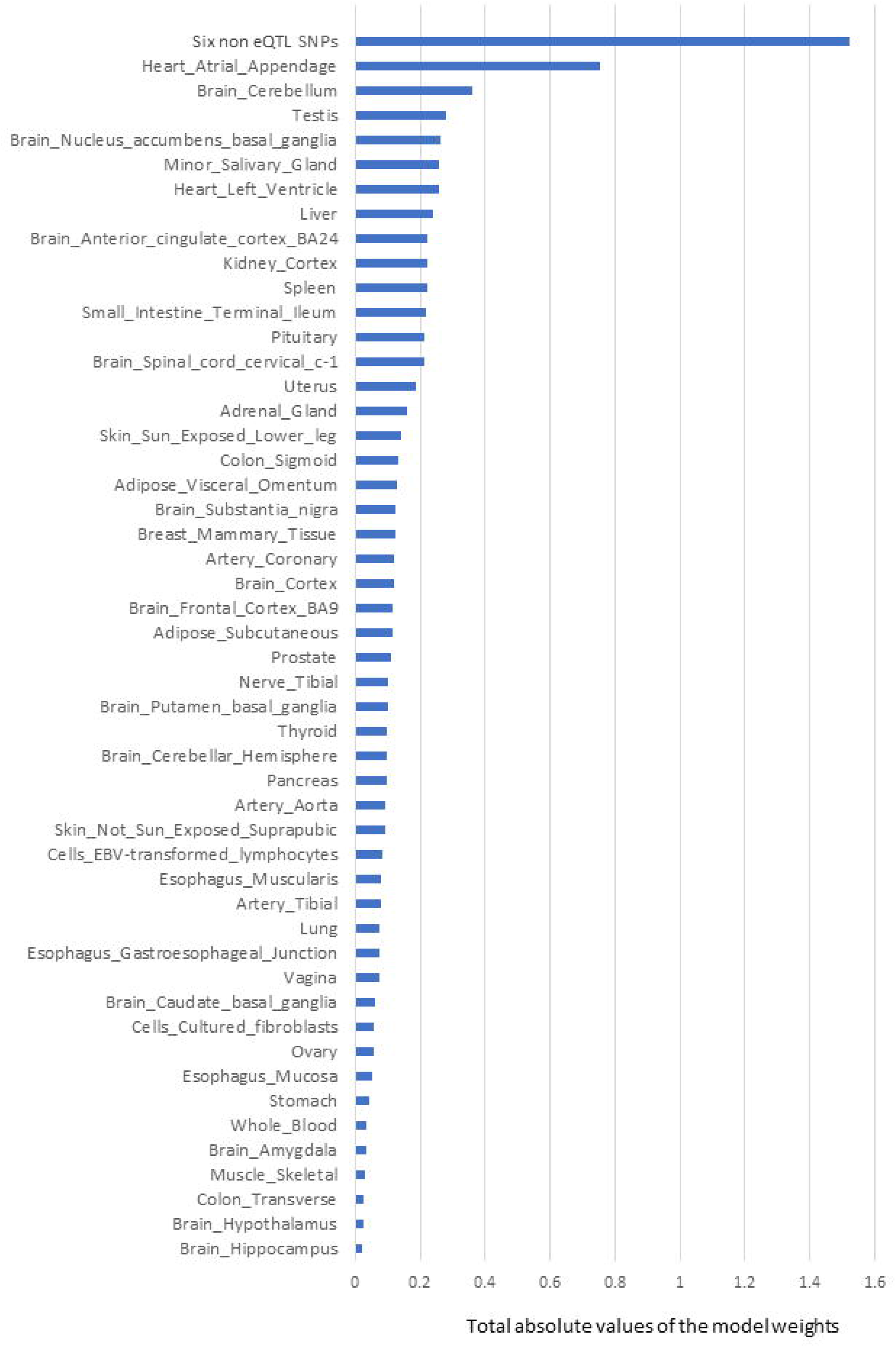
The rank order of tissue-specific risk contributions to risk of developing PD calculated using model-1. Tissue PD risk contributions were the sum of the absolute values of the model weights (coefficients) of the features used in the logistic regression predictor (model-1) according to their tissues. The SNPs/eQTLs that contributed to each category are listed (Table 1).

The next most significant contributions to the risk of PD development involved eQTLs that affected the heart atrial appendage (9%) and brain cerebellum (4%; Figure 2). The substantia nigra is viewed as a central brain region in PD yet eQTL gene regulation specific to the substantia nigra contributed ∼1.5% of the risk of PD development. We repeated the calculation of the tissue-specific contribution ranking using data from the 50 optimised predictor models, generated with model-1’s hyperparameters by 5 repeats of 10-fold cross validation (randomizing the full Mann-Whitney U test filtered PD variant derived eQTL effect matrix),. Again, SNPs lacking known eQTL effects, heart atrial appendage, and brain cerebellum were identified as the top three genetic contributors to the risk of PD development (Figure 3).

**Figure 3:**
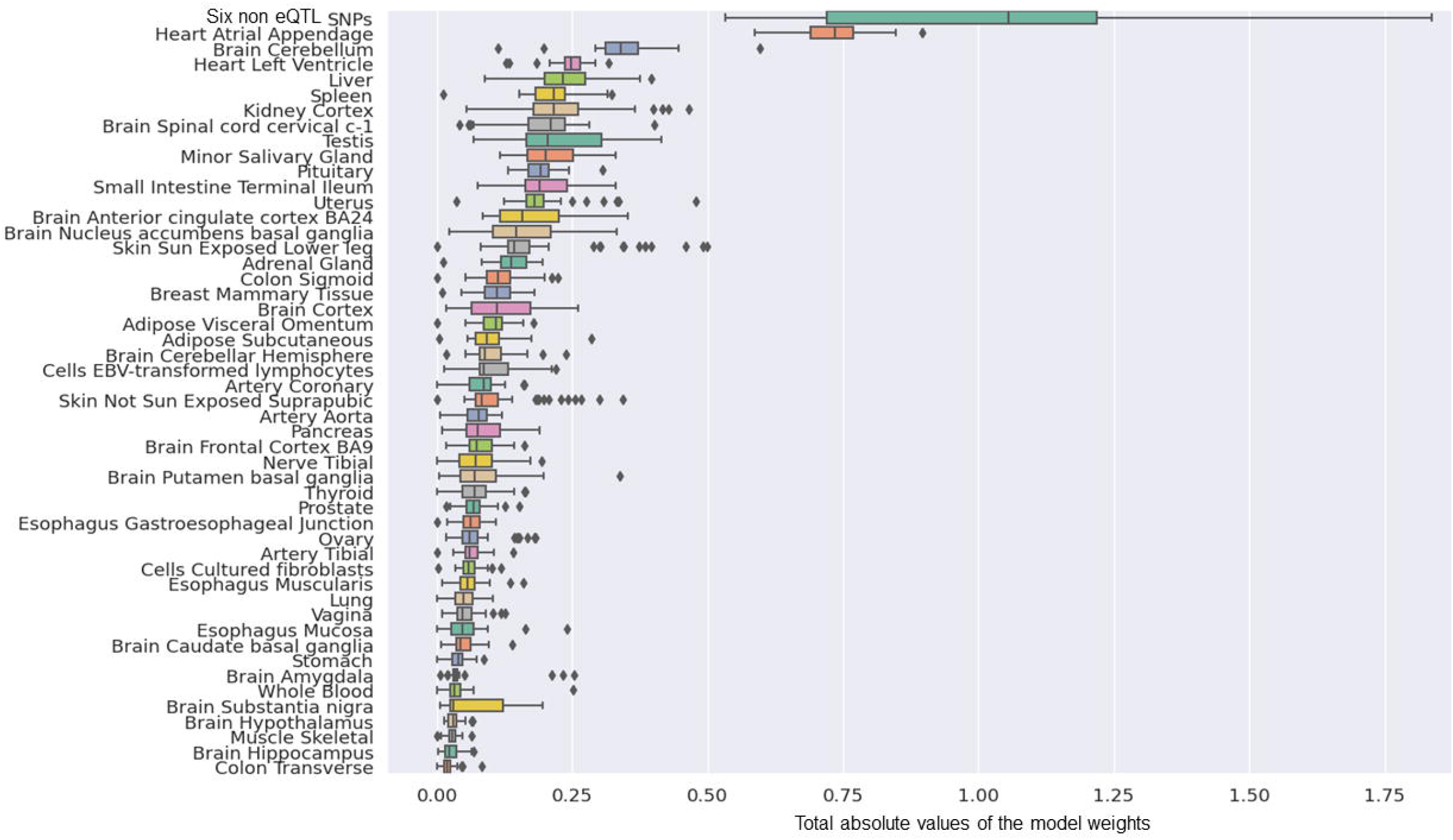
The rank order of tissue-specific risk contributions calculated across 50 predictor models created from randomised modelling and model-1’s hyperparameters. The tissue ranking was consistent with that observed for model-1.

Fifteen eQTLs contributed to the heart atrial appendages contribution to the risk of developing PD measured in model-1 (Table 1). Notably, the two biggest eQTL contributors, rs7617877 and rs6808178, each accounted for approximately 3% to the total model weight. rs7617877 and rs6808178 are in high linkage disequilibrium (R^2^ = 0.86)^39^ within European populations. rs7617877 and rs6808178 do not show detectable spatial regulatory associations with their nearest genes and instead both act as eQTLs for a gene > 13Mb downstream, *EAF1-AS1*, in the heart atrial appendage. *EAF1-AS1* is a long antisense non-coding RNA gene transcribed in antisense to *EAF1*, that undergoes an isoform switch, and has a significantly different transcript usage in the brains of patients with Parkinson’s disease^40^. Interestingly, rs6808178 also acts as a heart atrial appendage eQTL for *TMEM161B-AS1* (Table 1), which has also been implicated in neurodegeneration^41^.

### Creating a PD logistic regression predictor model using the 90 SNPs from the PRS calculated by *Nalls et al*

In the latest PD GWAS meta-analysis, *Nalls et al*. identified 90 SNPs that contribute to a PRS model for PD risk^2^. We therefore sought to understand the PD risk contribution that was specific to these 90 SNPs and created a logistic regression predictor model using only this subset. 88 of the 90 variants passed quality control (post-imputation data cleaning and quality checking). The 88 SNPs were integrated with the WTCCC PD genotype data to create a PD SNP derived eQTL effect matrix of WTCCC individual samples (4366 individual samples: 1698 cases and 2668 controls). The PD SNP derived eQTL effect matrix contained 3,206 features consisting of related tissue-specific eQTL-eGene pairs (76 SNPs, 518 genes, 49 tissue types) and 12 SNPs that lacked CoDeS3D detectable eQTL effects. Mann-Whitney U test filtering (FDR < 0.05) left 920 features (12 SNPs, 95 genes, 49 tissue types) that were used in the subsequent logistic regression modelling^26^. Model training was repeated using the optimised predictor hyperparameters and the eQTL effect matrix for the full WTCCC cohort to create predictor model-2. Model-2 achieved in-sample PD prediction with an AUC = 0.604 using 311 features (12 SNPs, 46 genes, 49 tissue types) (Supplementary Table 11) that included 308 tissue-specific eQTLs and 3 SNPs without known eQTL effects. Model-2 was validated using the UK Biobank^17^ (AUC = 0.554) and NeuroX-dbGap^16,29^ (AUC = 0.568) genotype data.

We determined the tissue-specific distribution for the 50 predictors that were created with model-2’s hyperparameters. The results we observed were consistent with what we observed using model-1 (Figure 4). Specifically, three SNPs (rs117896735, rs35749011 and rs5019538) with no identifiable eQTL effects (Table 2) and the eQTLs within the heart atrial appendage were the top contributors to the risk of developing PD (Figure 4 and Table 2). The three non-eQTL SNPs appeared in both Model-1 and Model-2 and were observed to have similar effect sizes (both magnitude and direction) across both models. Also consistent with model-1, model-2 identified rs6808178 as the top eQTL contributing to the heart atrial appendage signal.

**Table 2:** SNPs identified as being the main contributors to model-2. a) SNPs with no detected eQTL effects, and b) eQTL effects within the Heart Atrial Appendage. The model weight is the coefficient assigned to each variant or eQTL in the logistic regression predictor model-2.

| **SNPs**<br>**(no detected eQTLs)** | **model**<br>**weight** |
| --- | --- |
| rs117896735_A | 0.54172 |
| rs35749011_A | 0.47224 |
| rs5019538_G | -0.04028 |

b)
| **Tissue** | **eQTL**<br>**(rsID_major allele)** | **Gene** | **model**<br>**weight** |
| --- | --- | --- | --- |
| Heart_Atrial_Appendage | rs6808178_T | EAF1-AS1 | 0.53154 |
| Heart_Atrial_Appendage | rs26431_G | EIF3KP1 | 0.0079 |
| Heart_Atrial_Appendage | rs504594_A | HLA-DQA2 | -0.00333 |
| Heart_Atrial_Appendage | rs62053943_T | RP11-259G18.3 | -0.0014 |
| Heart_Atrial_Appendage | rs62053943_T | DND1P1 | -0.00092 |
| Heart_Atrial_Appendage | rs62053943_T | KANSL1-AS1 | -0.00064 |
| Heart_Atrial_Appendage | rs62053943_T | LRRC37A4P | 0.00061 |

**Figure 4:**
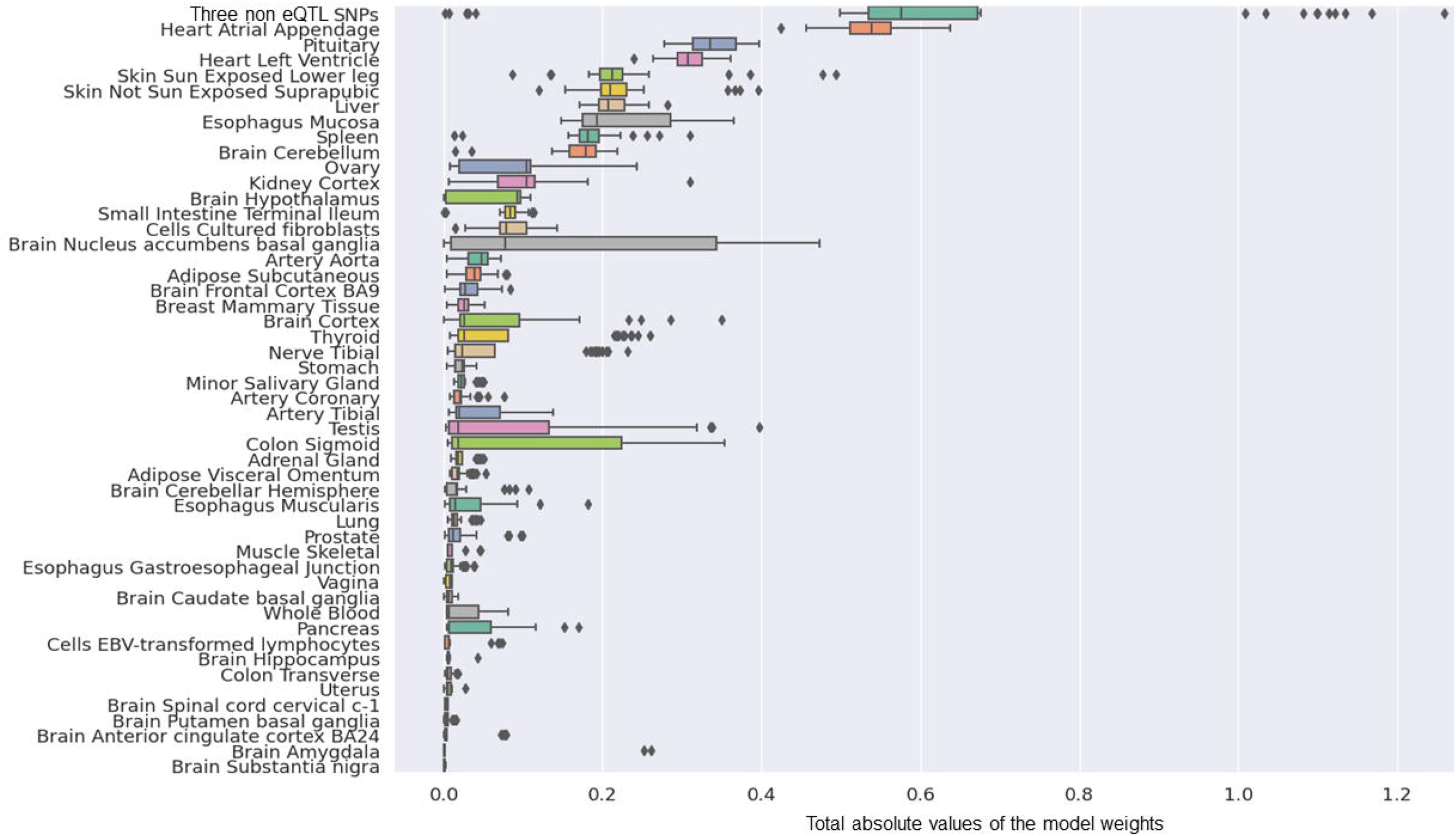
The group contributions of 50 predictors created with model 2 hyperparameters by 5 repeats of 10 fold cross-validation.

## Discussion

The mechanisms by which PD-associated genetic variants^4,16,42,43^ contribute to disease risk and development have not been fully elucidated. Yet, it is critical that we identify the mechanisms by which they impact on PD because this will allow patient stratification and the development of therapeutics that target disease progression and not just pathology. We used machine learning to understand the genetic architecture of PD risk, by identifying and ranking the pivotal risk variants and tissue-specific eQTL effects that contribute to such risk. Curated PD-associated SNPs from the GWAS catalogue^44^ were analysed to identify their tissue-specific eQTL effects. Regularised logistic regression predictor models that evaluated PD risk were built and validated across three independent case:control cohorts^3,16,17^. Model-1 achieved superior predictivity in comparison to Model-2, and delivered an in-sample predictive AUC = 0.627, and was subsequently validated in two independent test datasets derived from the UK Biobank^17^ (AUC = 0.572) and NeuroX-dbGap^16^ (AUC = 0.571). Although greater PRS predictivity has been achieved for PD by other groups, our main aim was to determine the SNPs-genes-tissue combinations that have the greatest contribution. Model-1 (generated from 290 SNPs) identified 6 SNPs without known eQTL effects and the SNP modulated gene regulation within the heart atrial appendage as being the major contributors to the predicted risk of developing PD. A second model (Model-2) that was generated using only 90 SNPs^2^ (which were previously identified to have the greatest predictive power with a PRS analysis) confirmed a subset of the top predictors we observed with model-1. Collectively, our results confirm roles for SNPs that are significantly connected with *INPP5P, CNTN1, GBA* and *SNCA* in PD and separately suggest a key role for transcriptional changes within the heart atrial appendage in the risk of developing PD. Effects associated with eQTLs located within the brain cerebellum were also recognized to confer major PD risk in the more extensive model (model-1) consistent with current hypotheses suggesting the brain cerebellum plays a role in PD development^45–47^.

For the top six contributing SNPs to the model, our analyses did not identify any spatial eQTL interactions. However, previous research has shown connections between these SNPs and three well-known PD-associated genes (*INPP5F, GBA, SNCA*)^48–51^, and an additional gene (*CNTN1*). rs117896735, the top contributor to model-1, is an intronic variant of *INPP5F* and has previously been identified as eQTL for *INPP5F* transcript levels (the IPDGC locus browser^52^). *INPP5F* is a known risk gene for PD^49^ that regulates STAT3 intracellular signalling pathways^53^ and has functional roles in cardiac myocytes and axons^54,55^. rs1442190 is an intronic variant within *CNTN1*, a known risk gene for dementia with Lewy bodies^56,57^ that encodes a cell adhesion protein, which is important for axon connections and nervous system development^58^. rs35749011 and rs12726330 are linked to the well-known PD-associated gene *GBA*^50^ through strong linkage disequilibrium connections (R^2^ = 0.77^39^) with rs2230288^50,59^, a missense coding variant located within *GBA*. rs35749011 has eQTL effects on *GBA* gene identified by the IPDGC database^52^. The final two SNPs, rs356220 and rs5019538, are located downstream of *SNCA. SNCA* encodes α-synuclein, which is central to PD pathogenesis^51^. The IPDGC database^52^ indicates that rs5019538 has eQTL effects on *SNCA*. Notably, rs356220 had the strongest association to PD in the original WTCCC GWAS^3^. Therefore, there is sufficient evidence that has previously associated these six variants with PD through connections to PD risk genes.

### Allele specific regulatory changes in the heart atrial appendage confer PD risk

We identified that eQTLs specific to the heart atrial appendage make a reproducible and substantial (2^nd^ highest) contribution to the risk of developing PD. The heart atrial appendage is a trigger site of atrial fibrillation (AF)^60^ and highly associated with hypertension and stroke^61–64^. There is a growing body of research indicating a close relationship between cardiovascular health and PD development^65–70^. For example, AF has been strongly related to early-stage PD^65^. Moreover, *Moon et al*. identified that patients with PD have an increased risk of AF, with a threefold increased risk (HR: 3.06, 95% CI: 1.20-7.77) of AF in younger PD patients (age: 40-49 years)^71^. Observations of a cross-sectional PD patient cohort have identified abnormal blood flow patterns in brains^72^ and it is argued that AF-associated perturbation of the brain blood supply networks promotes tissue inflammation and damage leading to PD pathogenesis^73^.

Amongst the 15 eQTL features that combined to make the heart atrial appendage’s contribution to the risk of developing PD (Table 1 & 2), eQTL up-regulation of *EAF1-AS1* (a long non-coding mRNA) made the greatest contribution. *EAF1-AS1* has different isoforms some of which overlap *EAF1* and *COLQ* (collagen like tail subunit of asymmetric acetylcholinesterase). Elevated *EAF1-AS1* transcript levels have previously been identified by differential gene expression analyses in brain tissue samples from PD patients^40^. It is interesting to speculate that the impact of this change is mediated through the interaction of *EAF1-AS1* with *EAF1*. Notably, *EAF1* has been associated with both neural development^74^ and TGF-β signalling^75^, which is a key pathway in many cardiac physiological processes^76^. As such, the deregulation of *EAF1-AS1* might impact on cardiac health. However, the anti-sense overlap is limited to the 3’ UTR of *EAF1* (UCSC Genome browser GRCh38/hg38). Therefore, we propose that future studies should investigate the regulatory impacts of *EAF1-AS1* on *EAF1* and the consequences of alterations in expression levels on heart function and PD disease. We contend that understanding this relationship may help to decipher the complex interactions connecting cardiovascular fitness and PD pathogenesis.

Similar to our work, *Li et al*.^77^ used linkage disequilibrium score regression (LDSC) analysis^78,79^ to identify enrichments of PD risk signals in six GTEx^14^ central nervous system tissues. However, three subsequent studies using LDSC have failed to reproduce *Li et al*.’s results ^80–82^. LDSC focuses on measuring the risk enrichment of genes uniquely expressed in each GTEx tissue^78,79^. By contrast, our model does not assume unique tissue expression. Rather, it identifies the risk associated with the PD-SNP, or the expression of all genes modulated specifically by PD-SNPs in different or multiple GTEx tissues. We therefore hypothesise that the fact that *Li et al*. did not identify any signals in heart tissues is likely due to the differences in the assumptions underlying the methodologies.

We acknowledge several limitations within our work. Firstly, the low predictive power of the models, in part, is due to the sample sizes and SNPs that were present within the cohorts we used to train and validate our models. We also acknowledge that the individuals in the included datasets are predominantly of European descent, and thus the significance of our findings are limited to this ethnicity. One limitation that impacts the vast majority of PD research is the lack of consistency in diagnostic criteria from one cohort to the other, and our study is not exempt from this.

The limitations within our study do not detract from the strengths of our model which included the fact that contributing features were: 1) easily identifiable; 2) validated across three independent cohorts; and 3) consistently identified genomic regions that are unanimously recognised as being associated with PD (*e*.*g. SNCA*).

Our approach provides a significant advance over other previously reported methods. The novelty revolves around the ability of our method to: 1) rank the contributions that SNPs make to a phenotype through regulatory changes; 2) identify the tissues in which these changes are occurring; and 3) include effects from variants that do not have detectable eQTLs in the reference library that is used in the assay. Finally, the consistency between models and ability to filter extraneous SNPs (e.g. T1D eQTLs) out of the final predictor is another strength of this study. The higher predictive power observed for model-1 (Supplementary Table 12) may be explained by the observation that the final model included more features (827 vs 308). However, given that model-1 leveraged 290 PD-associated SNPs, the result also suggests that the 90 SNPs, originally identified as part of the *Nalls et al*. PRS analysis^2^, do in fact contain the major genetic components that are associated with the risk of developing PD. Therefore, while other genetic signals clearly remain to be identified, the finding that both models consistently identified the same SNPs and heart atrial appendage eQTLs as the top contributors to the risk of developing PD further confirms the significance of these observations.

## Conclusion

In conclusion, we applied machine learning algorithms to rank the pivotal variants and tissue-specific eQTL effects that may contribute to the risk of developing PD by integrating PD-associated SNPs with information on genome organisation, tissue-specific eQTLs and the genotypes of PD cases and controls. Across our two models we consistently identified the same SNPs and heart atrial appendage eQTLs, linked to *EAF1-AS1* regulation, as the top contributors to the risk of developing PD. These findings warrant further mechanistic investigations to determine if they are early contributors to PD risk and disease development.

## Data Availability

The data that support the findings of this study are available from the following resources in the public domain:
The SNPs are listed in Supplementary table 1.
The HiC datasets are listed in Supplementary table 2.
eQTL information from 49 human tissues were obtained from the Genotype-Tissue Expression database [GTEx] v8; www.gtexportal.org
PD genotype datasets were acquired from the: Wellcome Trust Case Control Consortium (Request ID 10584); UKBioBank (Application Number 61507); NeuroX-dbGap (dbGap project#98581-1)

http://www.gtexportal.org

## Acknowledgements

The authors would like to thank Tayaza Fadason and the Genomics and Systems Biology Group at the Liggins Institute for their helpful discussions. The eQTL data used for the analyses were obtained from Genotype-Tissue Expression (GTEx) Portal. We would like to acknowledge the funders of GTEx Project – common Fund of the Office of the Director of the National Institutes of Health, and by NCI, NHGRI, NHLBI, NIDA, NIMH, and NINDS.

This study makes use of data generated by the Wellcome Trust Case-Control Consortium. A full list of the investigators who contributed to the generation of the data is available from www.wtccc.org.uk. Funding for the project was provided by the Wellcome Trust under award 076113, 085475 and 090355.

This research has been conducted using the UK Biobank Resource under Application Number 61507.

## Authors’ roles

DH was responsible for research project execution (1A), statistical analysis, design, execution (2A & B); manuscript preparation, writing of first draft (3A). WS, AKL, and JO co-supervise DH and were responsible for statistical analysis review and critique (2C). WS, AKL, AC, SF, JOS were responsible for manuscript review and critique (3B). JO was responsible for research project conception, organization and execution (1A, B and C).

## Financial Disclosures of all authors (for the preceding 12 months)

Full Financial Disclosures of all Authors for the Past Year: Information concerning all sources of financial support and funding for the preceding twelve months, regardless of relationship to current manuscript, must be submitted with the following categories suggested.

## References

1. Clarke, C. E. et al. UK Parkinson’s Disease Society Brain Bank Diagnostic Criteria. vol. Appendix 1 (NIHR Journals Library, 2016).

2. Nalls, M. A. et al. Identification of novel risk loci, causal insights, and heritable risk for Parkinson’s disease: a meta-analysis of genome-wide association studies. Lancet Neurol. 18, 1091–1102 (2019).

3. Spencer, C. C. A. et al. Dissection of the genetics of Parkinson’s disease identifies an additional association 5’ of SNCA and multiple associated haplotypes at 17q21. Hum. Mol. Genet. 20, 345–353 (2011).

4. Visscher, P. M. et al. 10 Years of GWAS Discovery: Biology, Function, and Translation. Am. J. Hum. Genet. 101, 5–22 (2017).

5. Visscher, P. M., Brown, M. A., McCarthy, M. I. & Yang, J. Five years of GWAS discovery. Am J Hum Genet 90, (2012).

6. Farrow, S. L. et al. Establishing gene regulatory networks from Parkinson’s disease risk loci. bioRxiv 2021.04.08.439080 (2021) doi:10.1101/2021.04.08.439080.

7. Fadason, T., Ekblad, C., Ingram, J. R., Schierding, W. S. & Justin, M. Physical Interactions and Expression Quantitative Traits Loci Identify Regulatory Connections for Obesity and Type 2 Diabetes Associated SNPs. Front. Genet. (2017) doi:10.3389/fgene.2017.00150.

8. Delaneau, O. et al. Chromatin three-dimensional interactions mediate genetic effects on gene expression. Science (80-.). 364, (2019).

9. Fadason, T., Schierding, W., Lumley, T. & O’Sullivan, J. M. Chromatin interactions and expression quantitative trait loci reveal genetic drivers of multimorbidities. Nat. Commun. 9, 1–13 (2018).

10. Yu, J., Hu, M. & Li, C. Joint analyses of multi-tissue Hi-C and eQTL data demonstrate close spatial proximity between eQTLs and their target genes. BMC Genet. 20, 43 (2019).

11. Duggal, G., Wang, H. & Kingsford, C. Higher-order chromatin domains link eQTLs with the expression of far-away genes. Nucleic Acids Res. 42, 87–96 (2014).

12. Aguet, F. et al. Genetic effects on gene expression across human tissues. Nature 550, 204–213 (2017).

13. Schierding, W. et al. Common Variants Coregulate Expression of GBA and Modifier Genes to Delay Parkinson’s Disease Onset. Mov. Disord. 35, 1346–1356 (2020).

14. Aguet, F. et al. Genetic effects on gene expression across human tissues. Nature 550, 204–213 (2017).

15. Ongen, H. et al. Estimating the causal tissues for complex traits and diseases. Nat. Genet. 49, (2017).

16. Nalls, M. A. et al. Large-scale meta-analysis of genome-wide association data identifies six new risk loci for Parkinson’s disease. Nat. Genet. 46, 989–993 (2014).

17. Bycroft, C. et al. The UK Biobank resource with deep phenotyping and genomic data. Nature 562, 203–209 (2018).

18. Siitonen, A. et al. Genetics of early-onset Parkinson’s disease in Finland: exome sequencing and genome-wide association study. Neurobiol. Aging 53, 195–e7 (2017).

19. Benjamini, Y. & Hochberg, Y. Controlling the false discovery rate: a practical and powerful approach to multiple testing. J. R. Stat. Soc. B 57, 289–300 (1995).

20. Purcell, S. et al. PLINK: a tool set for whole-genome association and population-based linkage analyses. Am. J. Hum. Genet. 81, 559–575 (2007).

21. Loh, P.-R. et al. Reference-based phasing using the Haplotype Reference Consortium panel. Nat. Genet. 48, 1443–1448 (2016).

22. Durbin, R. Efficient haplotype matching and storage using the positional Burrows-Wheeler transform (PBWT). Bioinformatics 30, 1266–1272 (2014).

23. McCarthy, S. et al. A reference panel of 64,976 haplotypes for genotype imputation. Nat. Genet. 48, 1279 (2016).

24. Christ, M., Braun, N., Neuffer, J. & Kempa-Liehr, A. W. Time Series FeatuRe Extraction on basis of Scalable Hypothesis tests (tsfresh – A Python package). Neurocomputing 307, 72–77 (2018).

25. Reiner, A., Yekutieli, D. & Benjamini, Y. Identifying differentially expressed genes using false discovery rate controlling procedures. Bioinformatics 19, 368–375 (2003).

26. Abraham, A. et al. Machine learning for neuroimaging with scikit-learn. Front. Neuroinform. 8, 14 (2014).

27. Vihinen, M. How to evaluate performance of prediction methods? Measures and their interpretation in variation effect analysis. BMC Genomics 13 Suppl 4, (2012).

28. Mehta, P. et al. A high-bias, low-variance introduction to Machine Learning for physicists. Phys. Rep. (2018) doi:10.1016/B978-0-08-044132-0.50011-3.

29. Nalls, M. A. et al. NeuroX, a fast and efficient genotyping platform for investigation of neurodegenerative diseases. Neurobiol. Aging 36, 1605.e7-1605.e12 (2015).

30. Auton, A. et al. A global reference for human genetic variation. Nature 526, 68–74 (2015).

31. de Leeuw, C. A., Mooij, J. M., Heskes, T. & Posthuma, D. MAGMA: Generalized Gene-Set Analysis of GWAS Data. PLoS Comput. Biol. 11, 1–19 (2015).

32. Pal, K., Forcato, M. & Ferrari, F. Hi-C analysis: from data generation to integration. Biophys. Rev. 11, 67–78 (2019).

33. Hi-c, D. et al. Mapping three-dimensional genome architecture through in situ DNase Hi-C. Nat Protoc. 11, 2104–2121 (2017).

34. Jiao, X. et al. DAVID-WS: A stateful web service to facilitate gene/protein list analysis. Bioinformatics 28, 1805–1806 (2012).

35. Burton, P. R. et al. Genome-wide association study of 14,000 cases of seven common diseases and 3,000 shared controls. Nature 447, 661–678 (2007).

36. McKnight, P. E. & Najab, J. Mann Whitney U Test. Corsini Encycl. Psychol. 1 (2010).

37. Wider, C. et al. Association of the MAPT locus with Parkinson’s disease. Eur. J. Neurol. 17, 483–486 (2010).

38. Tobin, J. E. et al. Haplotypes and gene expression implicate the MAPT region for Parkinson disease: The GenePD Study. Neurology 71, 28–34 (2008).

39. Machiela, M. J. & Chanock, S. J. LDlink: A web-based application for exploring population-specific haplotype structure and linking correlated alleles of possible functional variants. Bioinformatics 31, 3555–3557 (2015).

40. Dick, F. et al. Differential transcript usage in the Parkinson’s disease brain. PLoS Genet. 16, 1–24 (2020).

41. Boros, F. A., Maszlag-Török, R., Vécsei, L. & Klivényi, P. Increased level of NEAT1 long non-coding RNA is detectable in peripheral blood cells of patients with Parkinson’s disease. Brain Res. 1730, 146672 (2020).

42. Escott-Price, V. et al. Polygenic risk of Parkinson disease is correlated with disease age at onset. Ann. Neurol. 77, 582–591 (2015).

43. Lill, C. M. et al. Impact of Parkinson’s disease risk loci on age at onset. Mov. Disord. 30, 847–850 (2015).

44. MacArthur, J. et al. The new NHGRI-EBI Catalog of published genome-wide association studies (GWAS Catalog). Nucleic Acids Res. 45, D896–D901 (2017).

45. Riou, A. et al. Functional Role of the Cerebellum in Parkinson Disease: A PET Study. Neurology (2021) doi:10.1212/WNL.0000000000012036.

46. Seidel, K. et al. Involvement of the cerebellum in Parkinson disease and dementia with Lewy bodies. Ann. Neurol. 81, 898–903 (2017).

47. Wu, T. & Hallett, M. The cerebellum in Parkinson’s disease. Brain 136, 696–709 (2013).

48. Riboldi, G. M. & Di Fonzo, A. B. GBA, Gaucher disease, and Parkinson’s disease: from genetic to clinic to new therapeutic approaches. Cells 8, 364 (2019).

49. Cao, M., Park, D., Wu, Y. & De Camilli, P. Absence of Sac2/INPP5F enhances the phenotype of a Parkinson’s disease mutation of synaptojanin 1. Proc. Natl. Acad. Sci. U. S. A. 117, 12428–12434 (2020).

50. Berge-Seidl, V. et al. The GBA variant E326K is associated with Parkinson’s disease and explains a genome-wide association signal. Neurosci. Lett. 658, 48–52 (2017).

51. Siddiqui, I. J., Pervaiz, N. & Abbasi, A. A. The Parkinson Disease gene SNCA: Evolutionary and structural insights with pathological implication. Sci. Rep. 6, 1–11 (2016).

52. Grenn, F. P. et al. The Parkinson’s Disease Genome-Wide Association Study Locus Browser. Mov. Disord. 35, 2056–2067 (2020).

53. Kim, H. S., Li, A., Ahn, S., Song, H. & Zhang, W. Inositol Polyphosphate-5-Phosphatase F (INPP5F) inhibits STAT3 activity and suppresses gliomas tumorigenicity. Sci. Rep. 4, 1–10 (2014).

54. Zhu, W. et al. Inpp5f is a polyphosphoinositide phosphatase that regulates cardiac hypertrophic responsiveness. Circ. Res. 105, 1240–1247 (2009).

55. Zou, Y. et al. Gene-silencing screen for mammalian axon regeneration identifies Inpp5f (Sac2) as an endogenous suppressor of repair after spinal cord injury. J. Neurosci. 35, 10429–10439 (2015).

56. Chatterjee, M. et al. Contactin-1 is reduced in cerebrospinal fluid of parkinson’s disease patients and is present within lewy bodies. Biomolecules 10, 1–15 (2020).

57. Guerreiro, R. et al. Investigating the genetic architecture of dementia with Lewy bodies: a two-stage genome-wide association study. Lancet Neurol. 17, 64–74 (2018).

58. Anderson, C. et al. PLP1 and CNTN1 gene variation modulates the microstructure of human white matter in the corpus callosum. Brain Struct. Funct. 223, 3875–3887 (2018).

59. Mata, I. F. et al. Large-scale exploratory genetic analysis of cognitive impairment in Parkinson’s disease. Neurobiol. Aging 56, 211.e1-211.e7 (2017).

60. Di Biase, L. et al. Left atrial appendage: An underrecognized trigger site of atrial fibrillation. Circulation 122, 109–118 (2010).

61. Stöllberger, C., Schneider, B. & Finsterer, J. Elimination of the left atrial appendage to prevent stroke or embolism?: anatomic, physiologic, and pathophysiologic considerations. Chest 124, 2356–2362 (2003).

62. Hart, R. G. & Halperin, J. L. Atrial fibrillation and stroke: concepts and controversies. Stroke 32, 803–808 (2001).

63. Turagam, M. K. et al. Epicardial Left Atrial Appendage Exclusion Reduces Blood Pressure in Patients With Atrial Fibrillation and Hypertension. J. Am. Coll. Cardiol. 72, 1346–1353 (2018).

64. Du, W. et al. Large left atrial appendage predicts the ablation outcome in hypertensive patients with atrial fibrillation. J. Electrocardiol. 63, 139–144 (2020).

65. Hong, C. T., Chan, L., Wu, D., Chen, W. T. & Chien, L. N. Association between Parkinson’s disease and atrial fibrillation: A population-based study. Front. Neurol. 10, (2019).

66. Scorza, F. A., Fiorini, A. C., Scorza, C. A. & Finsterer, J. Cardiac abnormalities in Parkinson’s disease and Parkinsonism. J. Clin. Neurosci. 53, 1–5 (2018).

67. Potashkin, J. et al. Understanding the links between cardiovascular disease and Parkinson’s disease. Mov. Disord. 35, 55–74 (2020).

68. Awerbuch, G. I. & Sandyk, R. Autonomic functions in the early stages of parkinson’s disease. Int. J. Neurosci. 74, 9–16 (1994).

69. Ascherio, A. & Tanner, C. M. Use of antihypertensives and the risk of parkinson disease. Neurology 72, 578–579 (2009).

70. Fang, X. et al. Association of Levels of Physical Activity With Risk of Parkinson Disease: A Systematic Review and Meta-analysis. JAMA Netw. open 1, e182421 (2018).

71. Han, S. et al. Increased atrial fibrillation risk in Parkinson’s disease: A nationwide population-based study. Ann. Clin. Transl. Neurol. 8, 238–246 (2021).

72. Teune, L. K. et al. Parkinson’s disease-related perfusion and glucose metabolic brain patterns identified with PCASL-MRI and FDG-PET imaging. NeuroImage Clin. 5, 240–244 (2014).

73. Junejo, R. T., Lip, G. Y. H. & Fisher, J. P. Cerebrovascular Dysfunction in Atrial Fibrillation. Front. Physiol. 11, (2020).

74. Liu, J. X. et al. Eaf1 and Eaf2 negatively regulate canonical Wnt/β-catenin signaling. Dev. 140, 1067–1078 (2013).

75. Liu, J. X. et al. Transcriptional factors Eaf1/2 inhibit endoderm and mesoderm formation via suppressing TGF-β signaling. Biochim. Biophys. Acta - Gene Regul. Mech. 1860, 1103–1116 (2017).

76. Yousefi, F. et al. TGF-β and WNT signaling pathways in cardiac fibrosis: Non-coding RNAs come into focus. Cell Commun. Signal. 18, 1–16 (2020).

77. Li, Y. I., Wong, G., Humphrey, J. & Raj, T. Prioritizing Parkinson’s disease genes using population-scale transcriptomic data. Nat. Commun. 10, 1–10 (2019).

78. Finucane, H. K. et al. Heritability enrichment of specifically expressed genes identifies disease-relevant tissues and cell types. Nat. Genet. 50, 621–629 (2018).

79. Finucane, H. K. et al. Partitioning heritability by functional annotation using genome-wide association summary statistics. Nat. Genet. 47, 1228–1235 (2015).

80. Reynolds, R. H. et al. Moving beyond neurons: the role of cell type-specific gene regulation in Parkinson’s disease heritability. npj Park. Dis. 5, (2019).

81. Gagliano, S. A. et al. Genomics implicates adaptive and innate immunity in Alzheimer’s and Parkinson’s diseases. Ann. Clin. Transl. Neurol. 3, 924–933 (2016).

82. Bryois, J. et al. Genetic Identification of Cell Types Underlying Brain Complex Traits Yields Novel Insights Into the Etiology of Parkinson’s Disease. Nat Genet 52, 482– 493 (2021).

