## Supplementary tables for "Machine learning identifies six genetic variants and alterations in the Heart Atrial Appendage as key contributors to PD risk predictivity": List of Supplementary Tables.docx

Machine learning identifies genetic variants and changes in the Heart Atrial Appendage as making significant contributions to PD risk.

Daniel Ho^1^, William Schierding^1^, Sophie Farrow^1^, Antony A. Cooper^2, 3^, Andreas Kempa-Liehr^4*^, Justin M. O’Sullivan^1,5,6,7*^

**List of Supplementary Tables**

S. Table 1: 290 PD –associated SNPs used in this study

S. Table 2: Hi-C datasets used in this study

S. Table 3: PD Illumina SNPs and individuals recommended for exclusion by the WTCCC

S. Table 4: Summary of CoDeS3D outputs for significant eQTLs for 290 PD-associated SNPs

S. Table 5: List of data features (eQTLs and SNPs) that were used in the modelling process.

S. Table 6 T1D SNPs (n=313) and PD GWAS SNPs (n=290) used in this study

S. Table 7: CoDeS3D output for sig eQTLs for the 313 T1D SNPs

S. Table 8: GWAS catalog association for the PD SNPs (downloaded on 2020-08-27-Orphanet 2828-withChildTraits).

S. Table 9: David functional Annotation/Gene Ontology tables for the PD eGenes (n=1334).

S. Table 10: PD Model-1 data features and model weights

S. Table 11: PD Model-2 data features and model weights

S. Table 12: PD risk predictive performance comparison of model1 and model2
